# Diagnostic Accuracy of Mid-Upper Arm Circumference for Screening Maternal Malnutrition: A Community-based Cross-sectional Study from Eastern India

**DOI:** 10.64898/2026.09.20.26363198

**Authors:** Shailesh Sanjul Hembrom, Kumari Asha Kiran, Manisha Kujur, Mary Pushpa Murmu, Surendra Sahu, Prerna Anand

## Abstract

**Objective:** Maternal malnutrition, encompassing both undernutrition and overnutrition, can adversely affect maternal and foetal health and therefore requires effective nutritional assessment during antenatal care. Although body mass index (BMI) is commonly used for nutritional assessment, its application during pregnancy has limitations and may be difficult to implement in low-resource settings. Mid-upper arm circumference (MUAC) is a simple, inexpensive and easily interpretable anthropometric measure that may be suitable for community-based screening. This study aimed to evaluate the diagnostic accuracy and utility of MUAC for screening malnutrition among pregnant women in a community-based setting in eastern India.

**Design:** This is a secondary analysis of data from a cross-sectional study. Anthropometric measurements were obtained using standardised procedures, Nutritional status assessed by MUAC was compared with body mass index (BMI). Diagnostic performance was evaluated using sensitivity, specificity, accuracy, positive predictive value (PPV), negative predictive value (NPV), Cohen’s Kappa, simple linear regression and receiver operating characteristic (ROC) curve analysis.

**Setting:** Selected health sub-centres (Ayushman Arogya Mandir) under the rural field practice area of a medical college in Ranchi district of Jharkhand, a state in eastern India.

**Participant:** A total of 337 pregnant women were randomly selected from antenatal care registers and enrolled after obtaining informed consent. Women with chronic medical conditions, such as hypertension, diabetes mellitus and hypothyroidism were excluded.

**Result:** Based on BMI, 21.4% of participants were underweight, while MUAC classified 38.3% as having moderate malnutrition and 0.6% as having severe acute malnutrition. For detecting undernutrition, MUAC demonstrated 94.4% sensitivity, 70.3% specificity, 76.4% accuracy, 51.9% PPV and 97.4% NPV. Agreement between BMI and MUAC was moderate (Cohen’s Kappa=0.509, p<0.001). ROC analysis identified 22.8 cm as the optimal cut-off for undernutrition, and 24.8 cm for overnutrition.

**Conclusion:** MUAC may be a useful and effective screening tool for maternal malnutrition, particularly in community and low-resource settings where conventional anthropometric assessment may be difficult. Further studies are required to establish robust MUAC cut-offs across different populations and gestational ages.

**Key Points:** *What is already known on this topic:* Maternal malnutrition is associated with adverse maternal and foetal outcomes, but nutritional assessment during pregnancy remains challenging. Mid-upper arm circumference (MUAC) is simple and feasible in low-resource settings, but evidence on its diagnostic performance and appropriate cut-offs in pregnant women is limited.

*What this study adds:* This study provides evidence of a strong correlation between MUAC and body mass index (BMI), a fairly high accuracy of MUAC for detecting undernutrition and optimal cut-offs of MUAC for undernutrition and overnutrition in the population under study.

*How this study might affect research, practice or policy:* The findings of this study provide evidence about the diagnostic accuracy of MUAC for screening of malnutrition in pregnant women, which can lead to further studies for deriving optimal cut-offs of MUAC in different populations. It also provides a basis for the government to adopt MUAC for detection of malnutrition in low-resource and community settings.

## Introduction

Malnutrition is a state of imbalance in an individual’s intake of energy and/or nutrients. Consequently, it includes both undernutrition and overnutrition, denoting a lack or excess of energy or nutrients, respectively. ^[1]^ Although malnutrition is a concerning issue for individuals of all ages, it is a cause of increased attention when occurring in pregnant women. The nutritional requirements of an expecting mother do not just cater to her bodily needs but also provide crucial nutrients to the foetus which impact its growth and development not just inside the womb but also influence the early years of the offspring. ^[2]^ Hence, it is of utmost importance that pregnant women receive proper nutrition before and during their pregnancy and the monitoring of their nutritional status becomes a critical part of ante-natal care (ANC) services.

Communities all over the world are now facing the threat of double burden of malnutrition (DBM), the simultaneous existence of undernutrition and overnutrition in the same population. ^[3]^ The reasons for this are manifold, such as changes in social and cultural environments of people, their changing lifestyles and increased access to unhealthy packaged and highly processed food items, to what has been termed as an obesogenic environment. ^[4,5]^ Either of undernutrition or overnutrition can have a negative impact on the pregnant mother or the foetus or both. Some of the complications of maternal undernutrition include low birth weight, small for gestational age and preterm birth, among others, while the same for maternal overnutrition include conditions such as gestational diabetes and hypertension, pre-eclampsia, postpartum haemorrhage, large for gestational age, congenital malformations and others. ^[6]^ So it is of utmost importance that pregnant women be screened and assessed for both ends of the malnutrition spectrum.

The calculation of body mass index (BMI) following anthropometry is one of the most popular methods employed for assessment of nutritional status of an individual, although, it use for the assessment of nutritional status of pregnant women is not a widely accepted method. ^[7]^ While a few authorities such as the Ministry of Health and Family Welfare, Government of India permit its use before 20 weeks’ gestation ^[8,9]^, others such as the Institute of Medicine (IOM), USA recommend the measurement of weight gain during each trimester taking pre-pregnancy BMI as baseline ^[10]^, as a superior method to assess nutritional status. Moreover, calculating BMI requires standard equipment and procedures which may not be feasible in low resource settings.

Mid-upper arm circumference (MUAC) is a widely accepted screening tool for undernutrition in children under 5 years. ^[11]^ However, its use in adult population including pregnant women is limited to research studies with few experts recommending its implementation. The Food and Nutrition Technical Assistance Project (FANTA) of the USAID (United States Agency for International Development) recommends its use for classification of nutritional status in adults including pregnant women and has also specified cut-off for the same which are based on a Médecins sans Frontières Switzerland review of literature between 1995 and 2012. ^[12,13]^ MUAC as a screening tool has certain advantages such as ease of use and interpretation and a light and simple equipment which can be easily used in low resource and field settings.

Globally, approximately 170 million women (9.1%) are underweight, while three times as many women (610 million or 32.5%) are overweight. The mean BMI of women globally is 24.4 kg/m^2^, which is indicative of an increasing burden of overnutrition. In India, 18.7% women are underweight, while 24.0% women are overweight or obese. ^[14]^ In Jharkhand, 26.2% women are underweight, while 11.9% women are overweight. ^[15]^ These figures for pregnant women are lacking due to no set standards for assessing nutritional status during the antenatal period.

To cater to the needs of pregnant women, the Government of India has launched various initiatives such as the Reproductive, Maternal, Neonatal, Child, Adolescent Health and Nutrition (RMNCHA+N) strategy ^[16]^ and the POSHAN Abhiyan ^[17]^, but the impact of these initiatives on maternal malnutrition needs to be studied by quantifying its burden.

The objective of this study was to evaluate the role of mid-upper arm circumference (MUAC) as a screening tool for malnutrition in pregnant women.

## Methods

The present study is a secondary analysis of data obtained from a larger community-based cross-sectional study conducted at seven health subcenters (Ayushman Arogya Mandir) coming under the rural field practice area of a medical college in Ranchi district, Jharkhand. The primary findings describing the prevalence and determinants of maternal malnutrition have been published previously. ^[18]^ This study aims to address a different research question by evaluating the diagnostic accuracy of mid-upper arm circumference (MUAC) as a screening tool.

### Sample size

Sample size was calculated using Cochran’s formula, 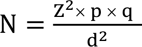, where Z = standard normal deviate (For 95% CI, the value was taken as 1.96), p = prevalence = 32% ^[11]^, q = (100-p) = 68%, d = allowable error (also called as precision) = 5%. The final sample size came out to be 334. For the purpose of the study, a total of 337 pregnant women were enrolled.

From each of the seven subcenters, 48 pregnant women were randomly selected from the antenatal care (ANC) registers and recruited after obtaining informed consent. Women who were willing to participate were eligible for inclusion, whereas those with chronic medical conditions such as hypertension, diabetes mellitus, or hypothyroidism, were excluded.

### Data Collection Procedure

Data were collected using a pre-tested, semi-structured questionnaire administered at the respective health subcenters. The questionnaire included items related to the variables under investigation. Interviews were conducted by the researcher in a comfortable setting after obtaining written informed consent form each participant. The researcher completed the questionnaire based on the participants’ responses.

The data collected included sociodemographic variables, and height, weight and mid-upper arm circumference (MUAC) of the study participants. The height was measured using a Seca 213 portable stadiometer ^[19]^, weight was measured using a Beurer PS 160 Digital Bathroom Scale ^[20]^ and MUAC was measured using an adult MUAC tape made up of non-tear stretch-resistant material as specified by UNICEF. ^[21]^ Body mass index was calculated and classified according to the guidelines provided by WHO. ^[1]^ Participants were classified into normal, moderate malnutrition and severe malnutrition on the basis of MUAC cutoffs as given by FANTA Project of USAID. ^[12,13]^

### Data Analysis

Data was entered into Microsoft Excel after generating a proper template. Data was analysed using Jamovi v 2.3. Sensitivity and specificity analysis, calculation of Cohen’s Kappa value, simple linear regression and ROC curve analysis was done to compare the findings of BMI and MUAC. BMI was chosen as the standard comparator despite its limitations during pregnancy due the limited time and resource setting of the study.

### Ethics approval

Study was conducted after approval by the Institutional Ethics Committee of RIMS, Ranchi as per Memo no. 10, IEC, RIMS Date 30.01.2023. Interviews with study subjects were conducted after obtaining their informed consent.

Patient and public involvement: Patients or the public were not involved in the design, or conduct, or reporting, or dissemination plans of our research.

## Results

### Socio-demographic profile of the study participants

The study included 337 pregnant women, with a mean age of 24.1 ± 3.61 years. Participants aged 18-25 years constituted the largest age group (70.6%). Regarding religion, 41.8% of the participants were Hindu. Tribal women accounted for 38.9% of the study population, while 61.1% belonged to the non-tribal group. In terms of educational attainment, matriculation was the most common level of education (31.5%). The majority of the participants were homemakers (88.7%). Based on the updated B.G. Prasad socioeconomic classification by 2024, the largest proportion of participants (40.9%) belonged to socioeconomic class IV (lower-middle class). ^[22]^ (Table 1)

**Table 1.** Socio-demographic characteristics of the study participants.

| **Socio-demographic variables** |  | **Frequency** | **Percentage** |
| --- | --- | --- | --- |
| Age group | 18-25 years | 238 | 70.6 |
|  | 26-30 years | 86 | 25.5 |
|  | 31-35 years | 12 | 3.6 |
|  | 36-40 years | 1 | 0.3 |
| Religion | Hindu | 141 | 41.8 |
|  | Sarna | 101 | 30 |
|  | Muslim | 93 | 27.6 |
|  | Christian | 2 | 0.6 |
| Ethnicity | Non-tribal | 206 | 61.1 |
|  | Tribal | 131 | 38.9 |
| Educational status | Post-graduate | 8 | 2.4 |
|  | Graduate | 30 | 8.9 |
|  | Intermediate | 94 | 27.9 |
|  | Matriculate | 106 | 31.5 |
|  | Up to high school | 85 | 25.2 |
|  | Literate without formal education | 8 | 2.4 |
|  | Illiterate | 6 | 1.8 |
| Occupation | Housewife | 299 | 88.7 |
|  | Student | 10 | 3 |
|  | Employed with private firm | 17 | 5 |
|  | Teacher | 6 | 1.8 |
|  | Farmer | 1 | 0.3 |
|  | Labor | 2 | 0.6 |
|  | PRI member | 1 | 0.3 |
|  | Anganwadi worker | 1 | 0.3 |
| Family type | Joint | 286 | 84.9 |
|  | Nuclear | 51 | 15.1 |
| Socio-economic status based on updated B.G. Prasad Classification Scale (2024) | Class I (Upper) | 8 | 2.4 |
|  | Class II (Upper middle) | 37 | 11 |
|  | Class III (Middle) | 121 | 35.9 |
|  | Class IV (Lower middle) | 138 | 40.9 |
|  | Class V (Lower) | 33 | 9.8 |

### Obstetric profile of the participants

The mean age at which the study participants had their menarche was 13±1.25 years. The mean gestational age at which the participants presented for the study was 17.5±8.43 weeks. Majority of the women had their menarche in normal time (88.7%), while nearly a tenth (9.8%) of the women had late menarche and a minor proportion (1.5%) achieved early menarche. Most of the participants (55.8%) were enrolled for the study during their second trimester, followed by first trimester (31.2%) and third trimester (13.1%). (Table 2)

**Table 2:**
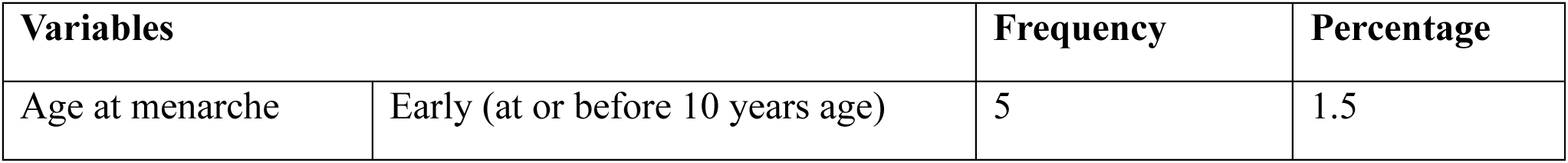
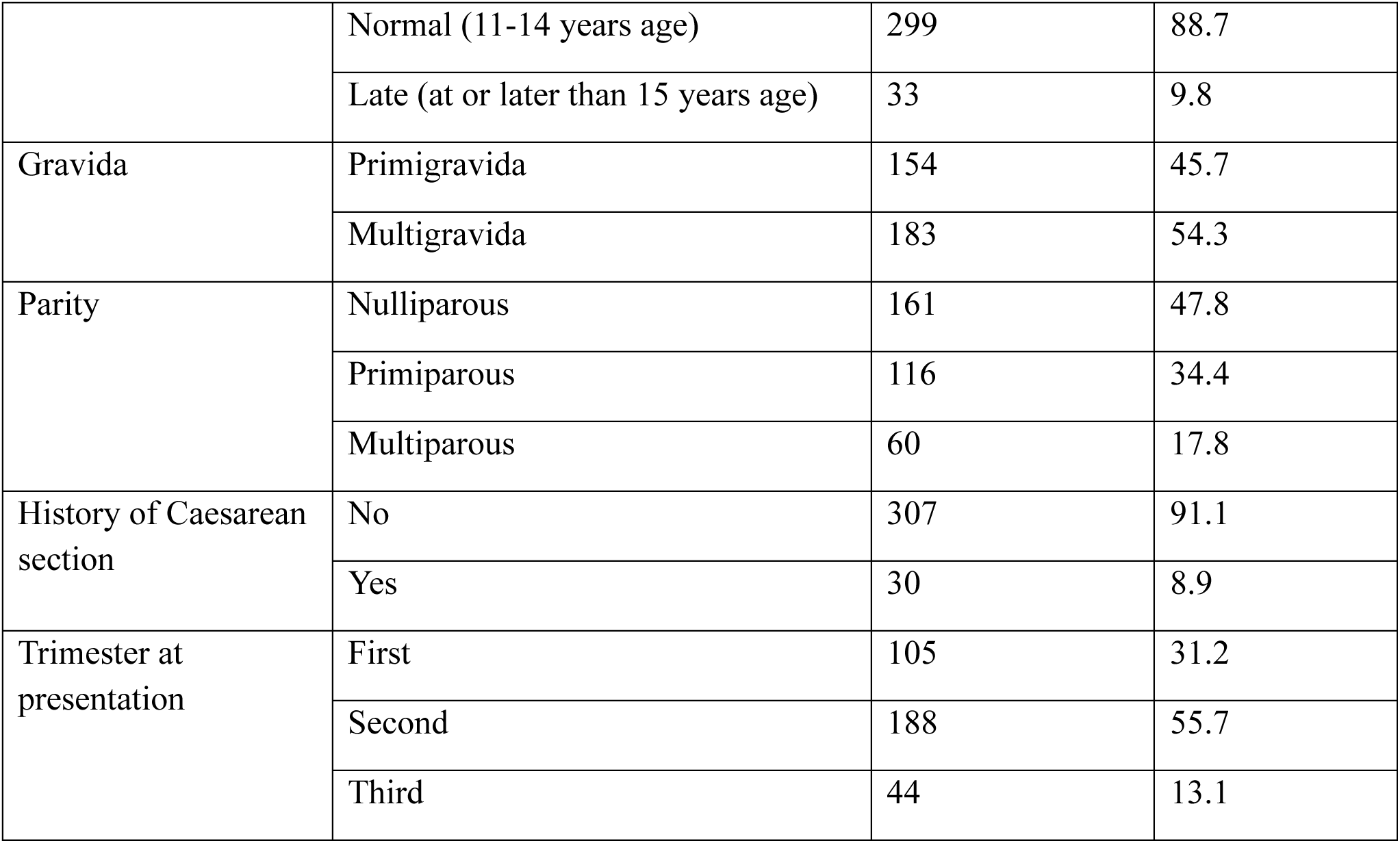
Obstetric profile of study participants (n=337)

### Nutritional status of study participants based on BMI

The mean values of height, weight and BMI of the participants were 1.51±0.05 meters, 48.7±8.3 kg, and 21.5±3.4 kg/m^2^ respectively. On the basis of Body Mass Index (BMI), 21.4% (72) of women were found to be underweight, while 14.8% (50) and 0.9% (3) women were found to be overweight and obese respectively. Nearly one-third (62.9%) of the women had a BMI within the normal range. (Table 2, Figure 1)

**Figure 1.**
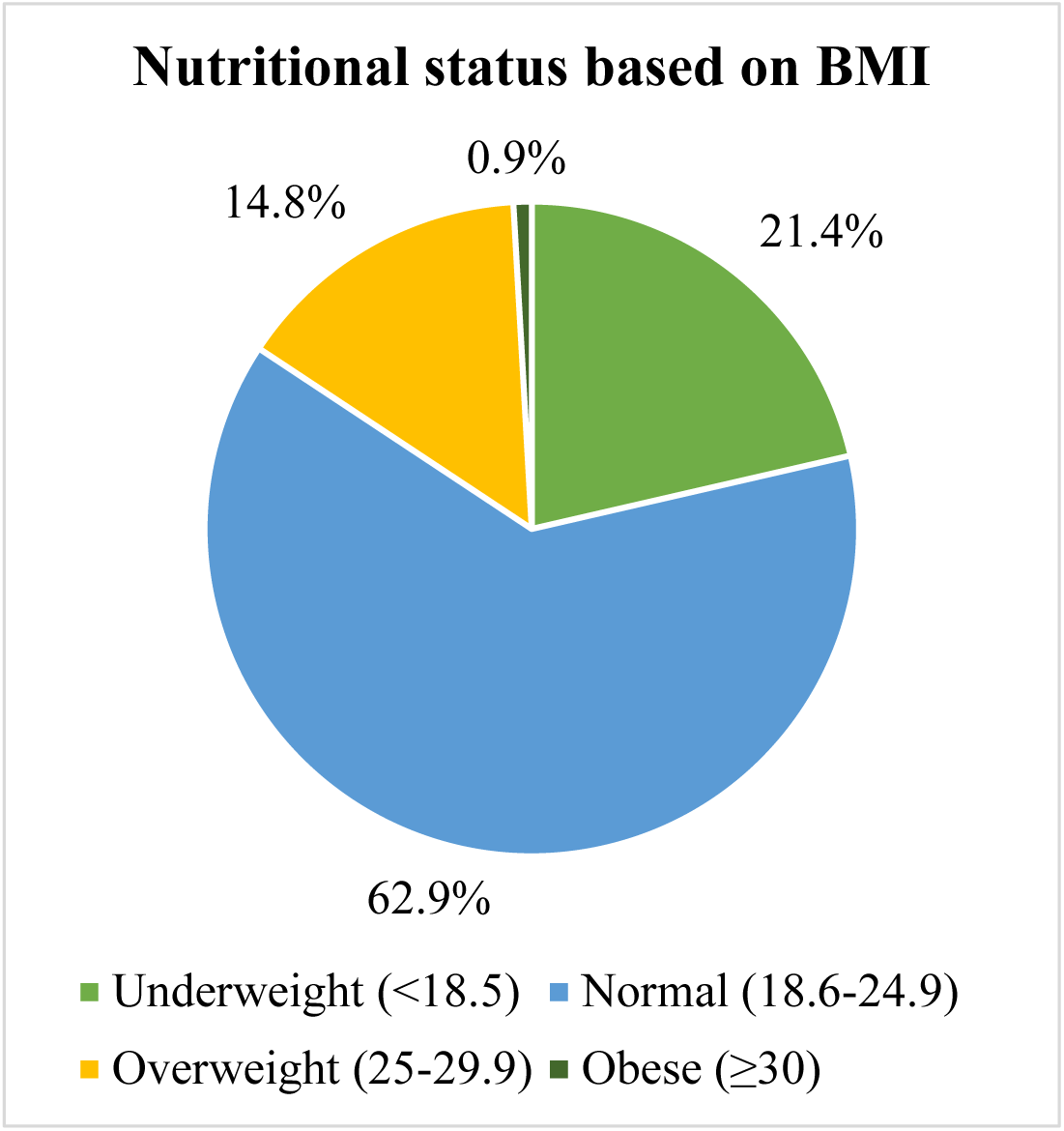
Nutritional status of participants based on BMI (n=337)

### Nutritional status of study participants based on MUAC

The mean MUAC value was 23.8±2.4 cm. On the basis of MUAC, 0.6% of women were found to be suffering from severe acute malnutrition, 38.3% from moderate malnutrition, while 61.1% women were found to be normal. (Table 3, Figure 2)

**Figure 2:**
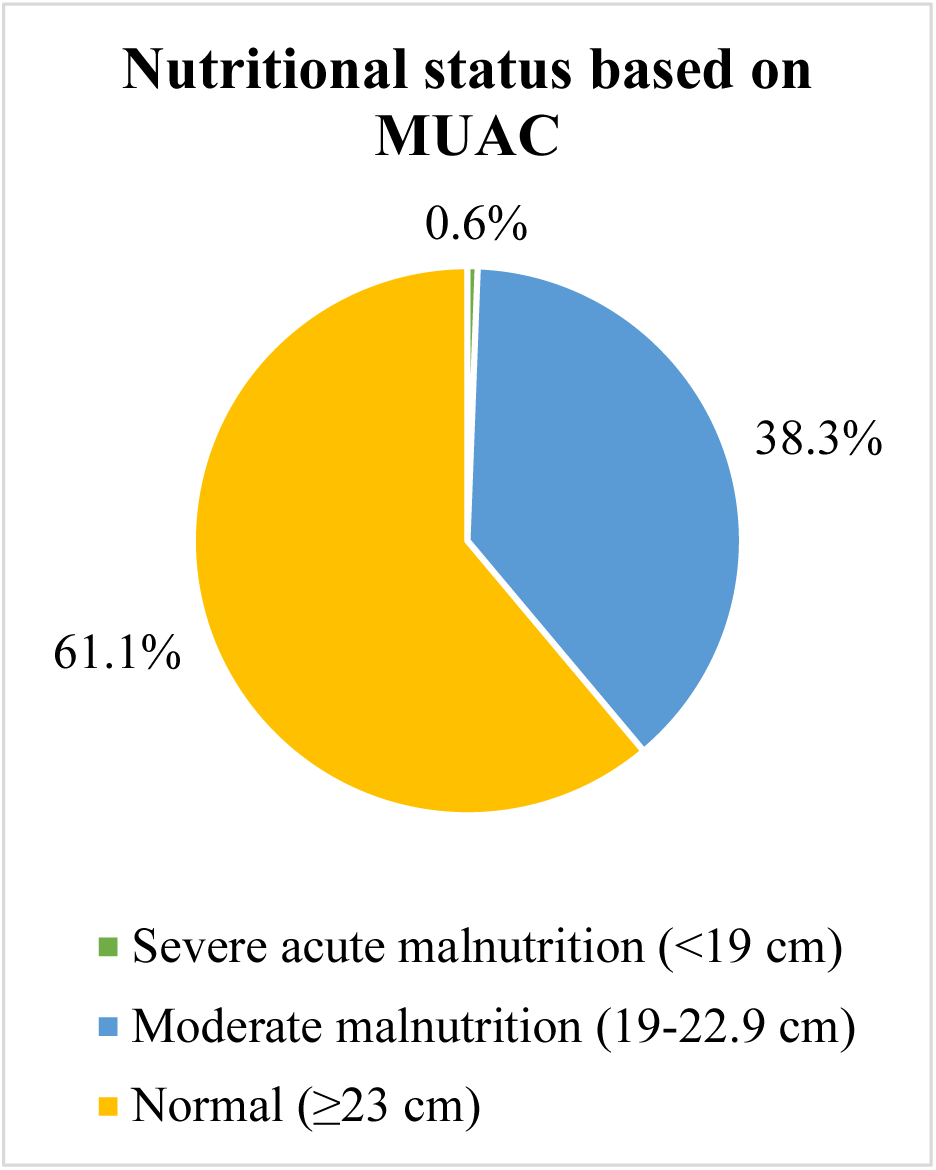
Nutritional status of participants based on MUAC (n=337)

**Table 3.**
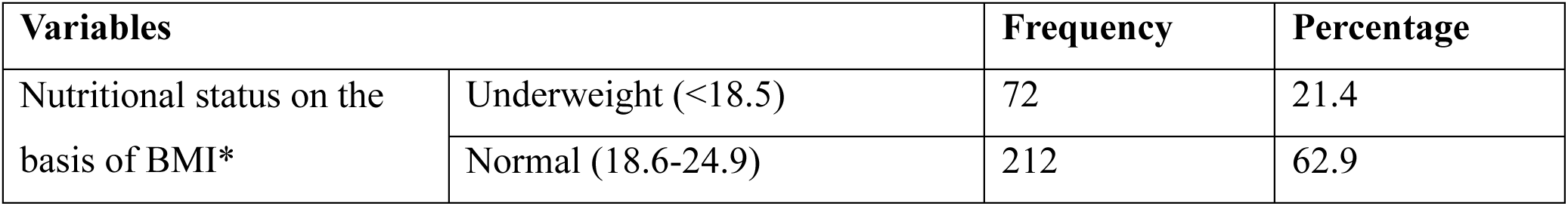
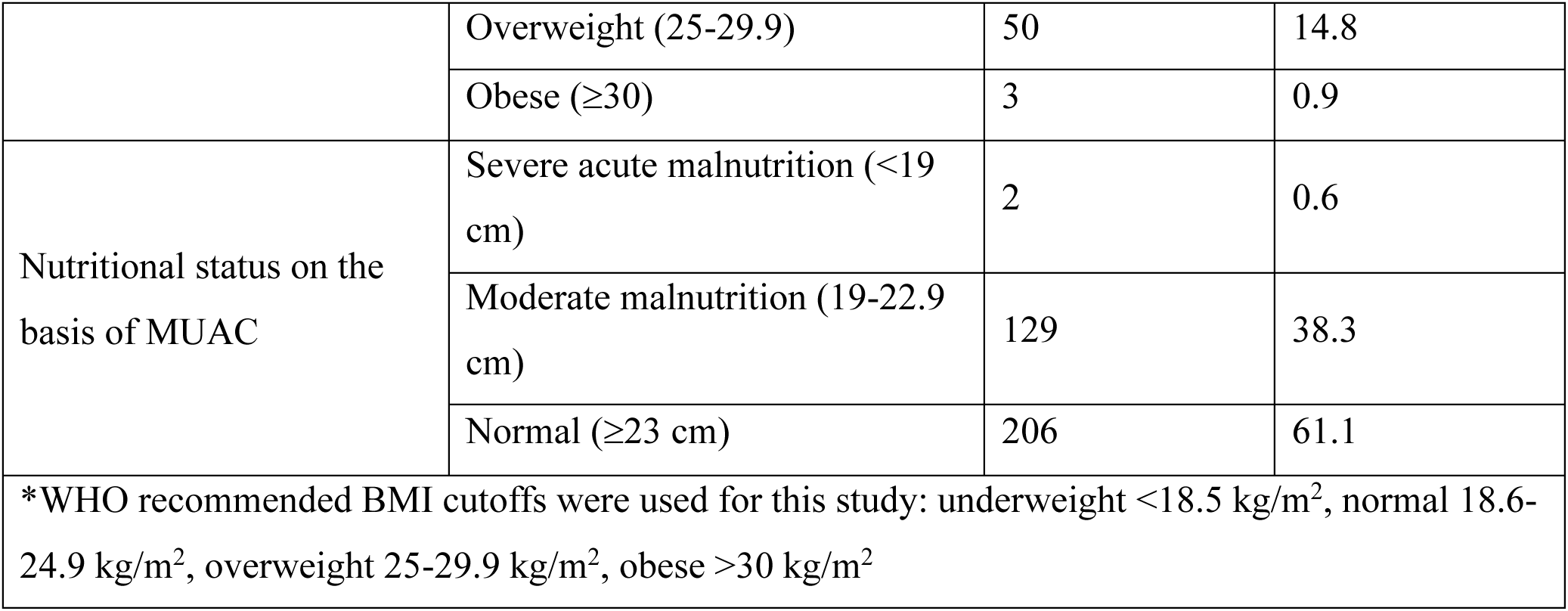
Classification of nutritional status of participants on the basis of BMI and MUAC (n=337)

### Comparison of results obtained by MUAC with BMI

On comparing the results of Mid-upper arm circumference (MUAC) with Body mass index (BMI) as a screening tool to identify undernutrition, MUAC had a sensitivity of 94.4%, specificity of 70.3%, accuracy of 76.4%, positive predictive value (PPV) of 51.9% and negative predictive value (NPV) of 97.4%. (Table 4)

**Table 4:** Comparison of nutritional status based on MUAC with BMI (n=284)

| Name of screening tool | Body mass index (BMI) |  |  |  |
| --- | --- | --- | --- | --- |
| Mid-upper arm circumference (MUAC) | Results of screening tool | Undernutrition | Normal | Total |
|  | Undernutrition | 68<br>True positive (TP) | 63<br>False positive (FP) | 131 |
|  | Normal | 4<br>False negative (FN) | 149<br>True negative (TN) | 153 |
|  | Total | 72 | 212 | 284 |

- Sensitivity = $\frac{\text{True positive}}{\text{True positive} + \text{False negative}} \times 100 = \frac{68}{68+4} \times 100 = 94.4\%$ - Specificity = $\frac{\text{True negative}}{\text{False positive} + \text{True negative}} \times 100 = \frac{149}{63+149} \times 100 = 70.3\%$ - Accuracy = $\frac{\text{True positive} + \text{True negative}}{\text{True positive} + \text{True negative} + \text{False positive} + \text{False negative}} \times 100 = \frac{68+149}{68+149+63+4} \times 100 = 76.4\%$ - Positive Predictive Value (PPV) = $\frac{\text{True positive}}{\text{True positive} + \text{False positive}} \times 100 = \frac{68}{68+63} \times 100 = 51.9\%$ - Negative Predictive Value (NPV) = $\frac{\text{True negative}}{\text{True negative} + \text{False negative}} \times 100 = \frac{149}{149+4} \times 100 = 97.4\%$

### Interrater reliability between BMI and MUAC for detecting undernutrition

For interrater reliability using Cohen’s Kappa, the agreement between BMI and MUAC for identifying undernutrition was 76% with a Kappa value of 0.509 indicating moderate agreement (p-value <0.001). (Table 5)

**Table 5.** Agreement between BMI and MUAC for identifying undernutrition (n=284)

| Screening result by BMI | Screening result by MUAC | Frequency | Percentage |
| --- | --- | --- | --- |
| Normal | Normal | 149 | 52.5 |
| Normal | Undernutrition | 63 | 22.9 |
| Undernutrition | Normal | 4 | 1.4 |
| Undernutrition | Undernutrition | 68 | 23.9 |

### Simple linear regression for relationship between BMI and MUAC

Simple linear regression was carried out to investigate the relationship between BMI (kg/m^2^) and MUAC (cm). The scatterplot showed that there was a strong positive linear relationship between the two, which was confirmed with a Pearson’s correlation coefficient of 0.790. Simple linear regression showed a significant relationship between BMI and MUAC (p < 0.001). The slope coefficient for MUAC was 1.10 so the BMI increases by 1.10 kg/m^2^ for each cm increase of MUAC. The R^2^ value was 0.624 so 62.4% of the variation in BMI can be explained by the model containing only MUAC. The relationship between MUAC and BMI can be shown by means of the equation BMI= −4.76 + 1.10 × (MUAC in cm). (Figure 3)

**Figure 3.**
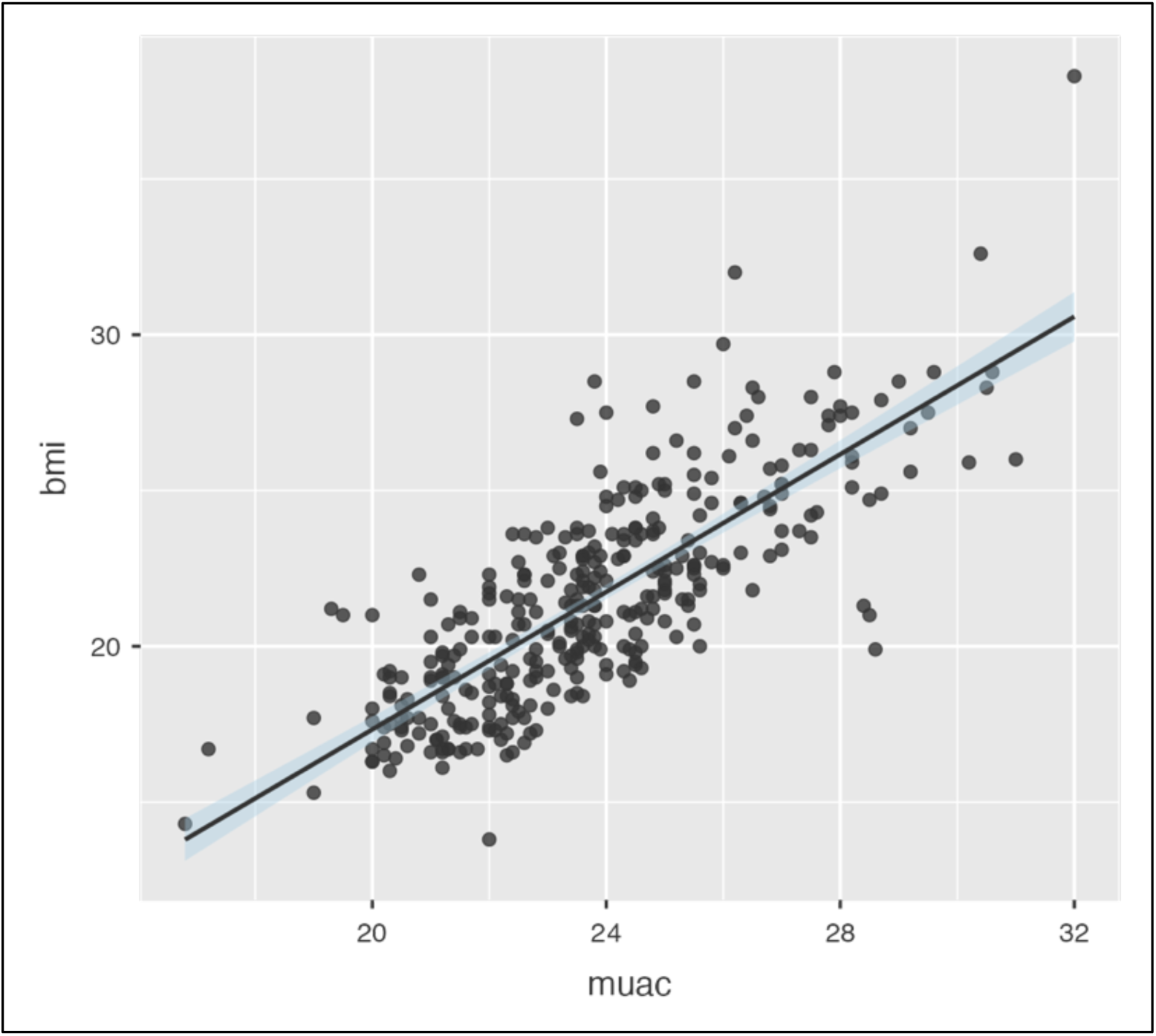
Scatterplot between MUAC (on X-axis) and BMI (on Y-axis) [n=337].

### ROC curve and AUC analysis for determining optimal cut-off points for undernutrition

A Receiver-Operating Characteristic curve analysis was done to determine the optimal cut-off values of MUAC to detect undernutrition among pregnant women. A cut-off value of 22.8 cm was revealed to be the optimal cut-off value with a sensitivity of 73.1%, specificity of 93.1%, and accuracy of 78.2%. The area under curve was 88.1% indicating an excellent diagnostic performance. The value of Youden’s index was 0.662 denoting the cut-off value as acceptable. (Table 6, Figure 4)

**Figure 4:**
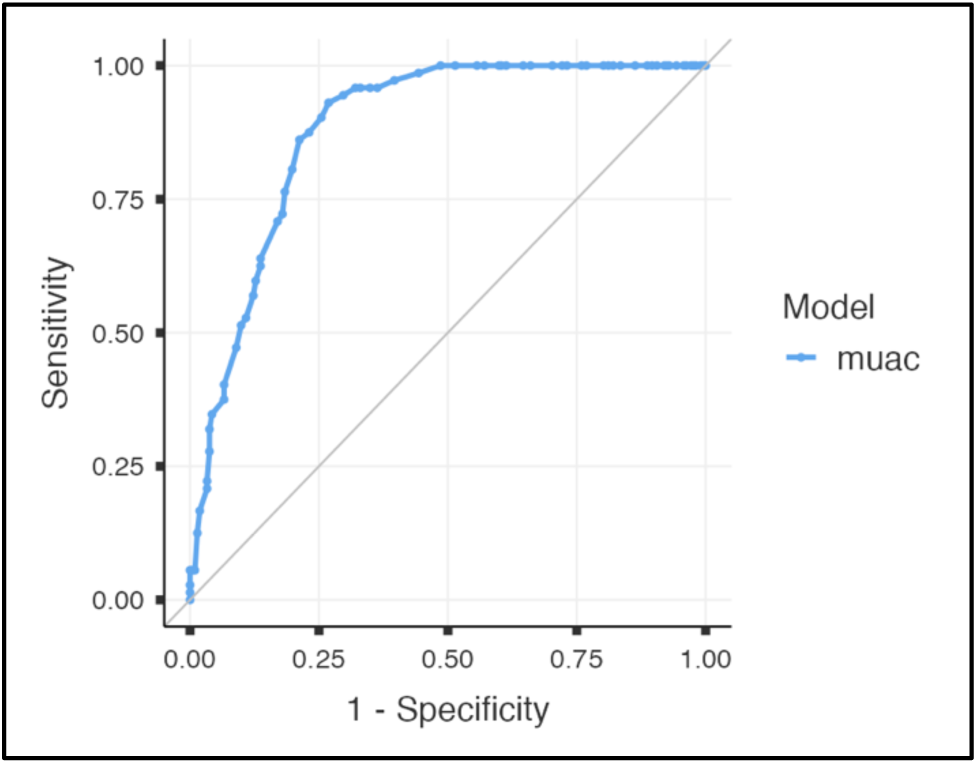
ROC curve of MUAC to detect undernutrition (n=284)

**Table 6.** Cut-off values of MUAC under ROC curve analysis for undernutrition and overnutrition.

| Cut-off (cm) | Sensitivity (%) | Specificity (%) | Accuracy (%) | PPV (%) | NPV (%) | Youden's index |
| --- | --- | --- | --- | --- | --- | --- |
| *Cut-off values in cm for undernutrition* |  |  |  |  |  |  |
| 19.1 | 5.6 | 100 | 76.1 | 100 | 75.7 | 0.056 |
| 22.4 | 93.1 | 78.8 | 80.6 | 57.9 | 94.4 | 0.649 |
| **22.8** | **93.1** | **73.1** | **78.2** | **54.0** | **96.9** | **0.662** |
| *Cut-off values in cm for overnutrition* |  |  |  |  |  |  |
| 23.9 | 96.2 | 55.7 | 63.8 | 35.2 | 98.3 | 0.519 |
| **24.8** | 86.8 | 73.1 | 75.8 | 44.7 | 95.7 | 0.599 |
| 25.4 | 75.5 | 83.5 | 81.9 | 53.3 | 93.2 | 0.590 |

### ROC curve and AUC analysis for determining optimal cut-off points for overnutrition

A Receiver-Operating Characteristic curve analysis was done to determine the optimal cut-off values of MUAC to detect overnutrition among pregnant women. A cut-off value of 24.8 cm was revealed to be the optimal cut-off value with a sensitivity of 86.79%, specificity of 73.11%, and accuracy of 75.85%. The area under curve was 88.6% indicating an excellent diagnostic performance. The value of Youden’s index was 0.599 denoting the cut-off value as acceptable. (Table 5, Figure 5)

**Figure 5:**
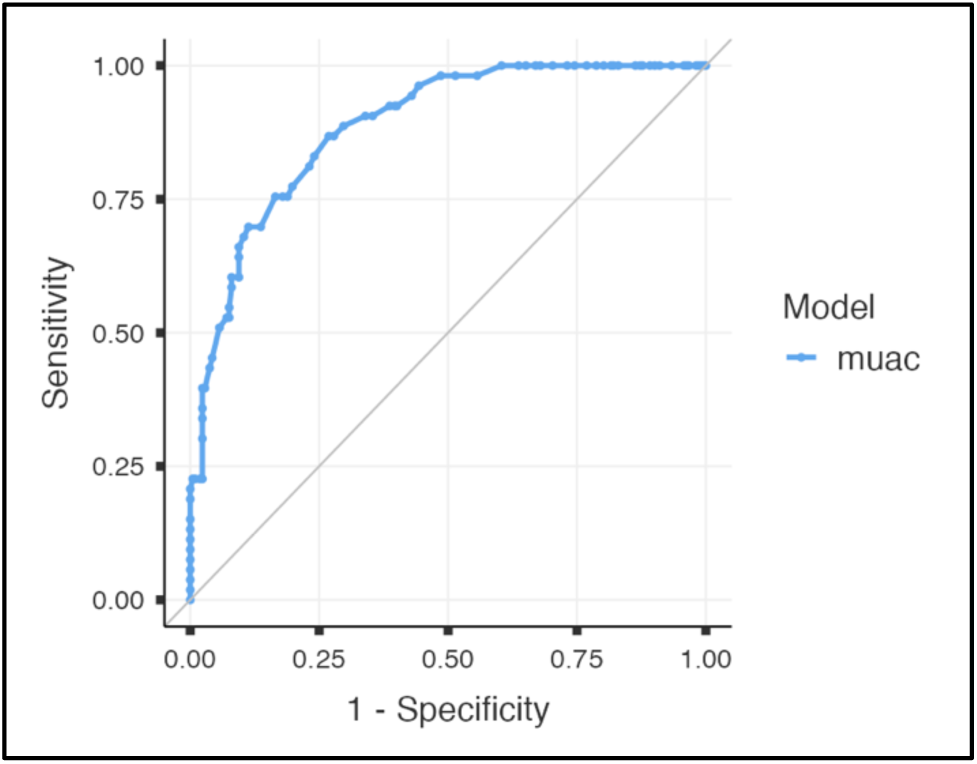
ROC curve of MUAC to detect overnutrition (n=265)

## Discussion

The findings and observations from our study revealed that 21.4% of women were found to be underweight on the basis of BMI, while this figure was 38.9% when MUAC was used. MUAC was found to be a fairly accurate tool for screening of undernutrition in pregnant women with a sensitivity of 94.4%, specificity of 70.3% and accuracy of 76.4%. There was moderate interrater agreement between MUAC and BMI for identifying undernutrition. Simple linear regression showed a significant and strong linear relationship between BMI and MUAC. The ROC curve analysis of MUAC determined cut-off values of 22.8 cm and 24.8 cm for detection of undernutrition and overnutrition among pregnant women.

Rathod et al. ^[23]^ reported mean height, weight and MUAC of 156.57 ± 4.460 cm, 57.104 ± 4.45 kg and 21.88 ± 2.260 cm, respectively, in a study conducted in tertiary care settings in Maharashtra in 2019. The proportion of undernutrition was 21.8%, taking a cut-off value of 21 com for MUAC. All parameters were better off than that in our study, which could be explained by the urban settings and the lower cut-off values taken.

A study Sen et al. ^[24]^ in 2020 reported an underweight proportion of 13.12% for MUAC cutoff value of 22 cm which was less than the 38.9% reported in our study. These findings may be reflective of the difference in settings of the two studies.

Saraswat et al. ^[25]^ reported prevalence of low MUAC (<23 cm) among pregnant women at 35.8% and 39.1% in Odisha and Chhattisgarh respectively in a study conducted in 2020. These findings do not coincide with those of NFHS-5, which reports lower rates of undernutrition on the basis of BMI in Chhattisgarh and Odisha compared to Jharkhand in women of reproductive age. This discrepancy could have arisen due to the fact that said study was conducted in a particular rural, tribal and poverty-stricken areas, while our study was done in a mixed rural and urban area.

In a prospective study done by Vasundhara et al. ^[26]^ on 928 pregnant women in 2020 in a tertiary hospital found 27.3% undernutrition based on MUAC cutoff of 22 cm. This is lower than that in our study (38.9%), which can be explained on the basis of the lower cutoff used and the urban setting of the study.

Mishra et al. ^[27]^ conducted a similar study in 2020 in Odisha and reported mean BMI and MUAC values of 21.81 ± 3.87 kg/m^2^ and 25.93 ± 2.76 cm, higher than our study. Taking Asian guidelines for BMI, 16.6% women were found to be underweight, while 29.1% women were found to be overweight, pre-obese or obese. The prevalence of undernutrition was higher in our study while the that for overnutrition could not be compared due to different cut-off values. The study considered a cut-off of 23.5 cm for MUAC and found 19.5% women to be underweight which was much lower than our study. This can be explained by the slightly different values for cut-off of MUAC and better nutritional status of women in general in Odisha compared to Jharkhand as evidenced by the NFHS-5 data.

A study conducted by Suresh et al. ^[28]^ in ANC clinics of urban areas of Delhi in 2021 found the mean height, weight and MUAC to be 152.15 ± 4.93 cm, 56.7 ± 6.7 kgs and 25.5 ± 2.60 cm respectively, which was higher than that in this study. 86% of women in the study had a normal MUAC (≥23 cm), which was higher than that found in this study at 61.1%. This difference can be attributed to the pre-existing anthropometric differences in women of reproductive age between the two regions as per NFHS-5 data.

The mean MUAC of normal weight women was 23.8 cm, that of underweight women was 21.3 cm and that of overweight women was 27.0 cm which is similar to the findings reported by Bhowmik et al. ^[29]^ in a study done among 498 early pregnant women (6-14 weeks) in Dhaka, Bangladesh in 2019.

The significant positive correlation between BMI and MUAC found in our study has also been reported by other studies ^[28,30–34]^, although there were geographical and ethnic variations in the cut-offs for detecting malnutrition.

### Strengths and Limitations

This study, being a community-based study and employing random sampling, captured a fairly representative sample of the population. Standard calibrated equipment was used to record anthropometric measurements. These recordings were conducted by a single observer, thus reducing inter-observer variation. Multiple methods were employed to assess the relationship between BMI and MUAC, namely, sensitivity analysis, interrater reliability, simple linear regression and ROC curve analysis.

Being a cross-sectional study, the study had limitations pertaining to temporal changes is anthropometric measurements during the period of pregnancy. Utilizing BMI as a tool to assess the nutritional status of pregnant women is not a widely accepted practice, with pre-pregnancy BMI and subsequent gestational weight gain being a more recommended method. The study failed to do a sub-group analysis in the different trimesters due to varying number of participants in all the three trimesters. Any confounding due to age, educational status, occupation, socio-economic status, gravida, parity, trimester at presentation and other factors was not addressed.

## Conclusion

Based on the findings of this study, it can be concluded that MUAC may be employed as a useful and effective tool for the screening of malnutrition in pregnant women. However, considering the limitations of this study, it is recommended that further studies be conducted to arrive at more robust cut-off values of MUAC in different populations.

## Data Availability

All data produced in the present study are available upon reasonable request to the authors.

## References

1. World Health Organization. Fact sheets - Malnutrition. https://www.who.int/news-room/fact-sheets/detail/malnutrition [Accessed 11th October 2022].

2. Dutta DC, Konar H. DC Dutta’s Textbook of Obstetrics: Including Perinatology and Contraception. 8th ed. New Delhi: Jaypee, The Health Sciences Publisher; 2015. P. 782.

3. The Lancet. The Double Burden of Malnutrition. https://www.thelancet.com/series-do/double-burden-malnutrition [Accessed 8th January 2026].

4. Powell P, Spears K, Rebori M. What is Obesogenic Environment? https://extension.unr.edu/publication.aspx?PubID=2810 [Accessed 8th January 2026].

5. Winichagoon P, Margetts BM. The double burden of malnutrition in low- and middle-income countries. In: Romieu I, Dossus L, Willett WC. Energy Balance and Obesity. Lyon (FR): International Agency for Research on Cancer; 2017. (IARC Working Group Reports, No. 10.) CHAPTER 2. https://www.ncbi.nlm.nih.gov/books/NBK565820/

6. Liu L, Ma Y, Wang N et al. Maternal body mass index and risk of neonatal adverse outcomes in China: a systematic review and meta-analysis. BMC Pregnancy Childbirth. 2019;19(1):105. doi:10.1186/s12884-019-2249-z PubMed PMID: 30922244; PubMed Central PMCID: PMC6440121.

7. McLaren S. Nutrition and Global Health. 1st ed. Wiley; 2022. https://onlinelibrary.wiley.com/doi/book/10.1002/9781394322596 doi:10.1002/9781394322596

8. National Health Systems Resource Centre. Care During Pregnancy and Childbirth Training Manual for CHO at AB-HWC. Ministry of Health & Family Welfare, Government of India; 2021. https://nhsrcindia.org/sites/default/files/2021-12/Care%20During%20Pregnancy%20and%20Childbirth%20Training%20Manual%20for%20CHO%20at%20AB-HWC.pdf [Accessed 10^th^ July 2024].

9. National Centre of Excellence and Advanced Research on Diets, Lady Irwin College. Strengthening Maternal Nutrition Services in Antenatal Care (8 contacts). Ministry of Health & Family Welfare, Government of India; 2020. http://nceard.roshni-cwcsa.co.in/UploadPDF/Document-2020-03-12-12-33-52.pdf [Accessed 10^th^ July 2024].

10. Institute of Medicine (US) Committee on Nutritional Status During Pregnancy and Lactation. Nutrition During Pregnancy: Part I Weight Gain: Part II Nutrient Supplements. Washington (DC): National Academies Press (US); 1990. 7, Energy Requirements, Energy Intake, and Associated Weight Gain during Pregnancy. https://www.ncbi.nlm.nih.gov/books/NBK235247/

11. UNICEF and The Ministry of Health. Mid-Upper Arm Circumference (MUAC) Tapes: A Simple Tool to Detect Child Wasting and Save Lives. Jakarta; 2023.

12. Food and Nutrition Technical Assistance III Project (FANTA). Nutrition Assessment, Counseling, and Support (NACS): A User’s Guide—Module 2: Nutrition Assessment and Classification, Version 2. U.S. Agency for International Development (USAID); 2016. https://www.fantaproject.org/sites/default/files/resources/NACS-Users-Guide-Module2-May2016.pdf

13. Ververs MT, Antierens A, Sackl A, et al. Which anthropometric indicators identify a pregnant woman as acutely malnourished and predict adverse birth outcomes in the humanitarian context? PLoS Curr. 2013;5: doi: 10.1371/currents.dis.54a8b618c1bc031ea140e3f2934599c8 PubMed PMID: 23787989; PubMed Central PMCID: PMC3682760.

14. United Nations Children’s Fund. UNICEF Programming Guidance. Prevention of malnutrition in women before and during pregnancy and while breastfeeding. New York: UNICEF; 2021.

15. International Institute for Population Sciences (IIPS) and ICF. National Family Health Survey-5 (NFHS-5), 2019-21. Mumbai, India: Ministry of Health and Family Welfare, Government of India.

16. Taneja G, Sridhar VSR, Mohanty JS, et al. India’s RMNCH+A Strategy: approach, learnings and limitations. BMJ Glob Health. 2019;4(3):e001162. doi:10.1136/bmjgh-2018-001162

17. Press Information Bureau, Government of India. Mission Poshan 2.0 Strengthening India’s Nutrition Ecosystem. https://www.pib.gov.in/www.pib.gov.in/Pressreleaseshare.aspx?PRID=2109222 [Accessed 20^th^ May 2026].

18. Hembrom SS, Kujur M, Sagar V, et al. Double Burden of Malnutrition Among Pregnant Women in Rural Jharkhand: Evidence from a Cross-Sectional Study. Cureus. 2024. doi:10.7759/cureus.74692

19. Seca. Seca 213 - Portable stadiometer. https://www.seca.com/en_us/products/all-products/product-details/seca213.html [Accessed 20th May 2026].

20. GmbH B. Beurer. Personal bathroom scale PS 160. Available from: https://www.beurer.com/global/p/72530/ [Accessed 20th May 2026].

21. UNICEF. Product Specifiication Sheet - Adult MUAC Tape. 2020. https://www.unicef.org/supply/media/3996/file/MUAC-tape-adult-specifications-May2020.pdf

22. K J A, Mishra A, Borle AL. Updated B. G. Prasad Scale for Socioeconomic Status Classification for the Year 2024. Indian J Pediatr. 2024;91(6):643–643. doi:10.1007/s12098-024-05131-z

23. Rathod MS, Borde AN, Patil SP, et al. Undernutrition and its association with socio-demographic factors among pregnant women attending tertiary health care hospital in northern Maharashtra: a cross-sectional study. Int J Community Med Public Health. 2019;6(10):4456. doi:10.18203/2394-6040.ijcmph20194512

24. Sen J, Roy A, Mondal N. Association of Maternal Nutritional Status, Body Composition and Socio-economic Variables with Low Birth Weight in India. Journal of Tropical Pediatrics. 2010;56(4):254–9. doi:10.1093/tropej/fmp102

25. Saraswat A, Unisa S, Reshmi RS, et al. Assessment of Nutritional Status of Pregnant Women based on Mid-upper Arm Circumference (MUAC) and Associated Factors in Poverty Pockets of Eastern India. The Journal of Family Welfare; 64: Special Issue 2019-20.

26. Vasundhara D, Hemalatha R, Sharma S, et al. Maternal MUAC and fetal outcome in an Indian tertiary care hospital: A prospective observational study. Matern Child Nutr. 2020;16(2):e12902. doi:10.1111/mcn.12902 PubMed PMID: 31833195; PubMed Central PMCID: PMC7083480.

27. Mishra KG, Bhatia V, Nayak R. Association between mid-upper arm circumference and body mass index in pregnant women to assess their nutritional status. Journal of Family Medicine and Primary Care. 2020;9(7):3321–7. doi:10.4103/jfmpc.jfmpc_57_20

28. Suresh M, Jain S, Kaul N. Evaluation of MUAC as a tool for assessing nutritional status during pregnancy (>20 weeks of gestation) in Delhi India. World Nutrition. 2021. 12(1), 65–72. 10.26596/wn.202112165-72

29. Bhowmik B, Siddique T, Majumder A, et al. Maternal BMI and nutritional status in early pregnancy and its impact on neonatal outcomes at birth in Bangladesh. BMC Pregnancy Childbirth. 2019;19(1):413. doi:10.1186/s12884-019-2571-5

30. Miele MJ, Souza RT, Calderon I, et al. Proposal of MUAC as a fast tool to monitor pregnancy nutritional status: results from a cohort study in Brazil. BMJ Open. 2021;11(5):e047463. doi:10.1136/bmjopen-2020-047463 PubMed PMID: 34031116; PubMed Central PMCID: PMC8149442.

31. Ali MH, AlHabardi N, Adam I. Assessment of mid-upper arm circumference for detecting obesity in pregnant women: a cross-sectional study. Front Glob Women’s Health. 2025;6:1554068. doi:10.3389/fgwh.2025.1554068

32. Salih Y, Omar SM, AlHabardi N, Adam I. The Mid-Upper Arm Circumference as a Substitute for Body Mass Index in the Assessment of Nutritional Status among Pregnant Women: A Cross-Sectional Study. Medicina (Kaunas). 2023 May 23;59(6):1001. doi: 10.3390/medicina59061001. PMID: 37374205; PMCID: PMC10304801.

33. Fakier A, Petro G, Fawcus S. Mid-upper arm circumference: A surrogate for body mass index in pregnant women. S Afr Med J. 2017;107(7):606. doi:10.7196/SAMJ.2017.v107i7.12255

34. Cooley SM, Donnelly JC, Walsh T, et al. The relationship between body mass index and mid-arm circumference in a pregnant population. Journal of Obstetrics and Gynaecology. 2011;31(7):594–6. doi:10.3109/01443615.2011.597892

